# A qualitative study of how maternal morbidities impact women’s quality of life during pregnancy and postpartum in five countries in sub-Saharan Africa and South Asia

**DOI:** 10.1101/2025.01.14.25320557

**Authors:** Martha Abdulai, Priyanka Adhikary, Sasha G. Baumann, Muslima Ejaz, Jenifer Oviya Priya, M. Bridget Spelke, Victor Akelo, Kwaku Poku Asante, Bitanya M. Berhane, Shruti Bisht, Ellen Boamah-Kaali, Gabriela Diaz-Guzman, Anne George Cherian, Zahra Hoodbhoy, Margaret P. Kasaro, Amna Khan, Janae Kuttamperoor, Dorothy Lall, Gifta Priya Manohari, Sarmila Mazumder, Karen McDonnell, Mahya Mehrihajmir, Wilbroad Mutale, Winnie K. Mwebia, Imran Nisar, Kennedy Ochola, Peter Otieno, Gregory Ouma, Piya Patel, Winifreda Phiri, Neeraj Sharma, Emily R. Smith, Charlotte Tawiah, Natalie J. Vallone, Allison C. Sylvetsky

## Abstract

**Aim:** Maternal morbidities present a major burden to the health and well-being of childbearing women. However, their impacts on women’s quality of life (QoL) are not well understood. This work aims to describe the extent to which the morbidities women experience during pregnancy and postpartum affect their QoL and identify any protective or risk factors.

**Methods:** This qualitative study included pregnant and postpartum women in Kenya, Ghana, Zambia, Pakistan, and India. Data were collected between November 2023 and June 2024. Participants were selected via purposive sampling, with consideration of age, trimester, and time since delivery. A total of 23 focus group discussions with 118 pregnant and 88 late (≥6 months) postpartum participants and 48 in-depth interviews with early (*≤*6 weeks) postpartum participants were conducted using semi-structured guides developed by the research team. Data was analyzed using a collaborative inductive thematic approach.

**Results:** Four overarching themes were identified across pregnancy and the postpartum period: (1) physical and emotional challenges pose a barrier to daily activities; (2) lack of social support detracts from women’s QoL; (3) receipt of social support mitigates adverse impacts of pregnancy and postpartum challenges on QoL; and (4) economic challenges exacerbate declines in women’s QoL during pregnancy and postpartum.

**Conclusions:** Bodily discomfort and fatigue were near-universal experiences. Physical and emotional morbidities related to childbearing limited women’s ability to complete daily tasks and adversely impacted their perceived QoL. Social and financial support from the baby’s father, family and/or in-laws, community members, and healthcare providers are important to mitigate the impacts of pregnancy and postpartum challenges on women’s health and well-being.

## INTRODUCTION

Despite recent global advancements, maternal mortality and morbidity remain a serious public health concern in low- and middle-income countries (LMICs) [1]. In 2020, the estimated maternal mortality ratio (MMR) in Africa was 531 deaths per 100,000 live births, accounting for 69% of maternal deaths worldwide and 117 per 100,000 live births in South East Asia [2, 3]. Maternal deaths are often referred to as the ‘tip of the iceberg’—whereas for every maternal death there are an estimated 50 to 100 cases of severe maternal morbidity—and there is a renewed global push for interventions that go beyond averting death [4–6]. Recent evidence from the Alliance for Maternal and Newborn Health Improvement (AMANHI) cohort study, in eight LMICs, found that one in three pregnant women experienced at least one direct maternal morbidity, with the burden twice as high for women in South Asia compared to sub-Saharan Africa [7]. Other studies, such as an observational study in Maharashtra, India, report maternal morbidity incidence rates exceeding 50%, with most complications occurring in the postpartum period [8].

Definitions of maternal morbidity are broad and most morbidity data comes from hospital-based studies, capturing only women who seek medical care for the morbidities they experience. The World Health Organization (WHO) defines maternal morbidity as “any health condition attributed to and/or complicating pregnancy and childbirth that has a negative impact on the woman’s well-being and/or functioning” [4]. Morbidities are further understood as direct (i.e. obstetric complications resulting from pregnancy or its management), indirect (i.e. existing conditions aggravated by pregnancy), and psychological (e.g. postpartum depression, attempted suicide) [9]. Complications contributing to morbidity vary in their severity—ranging from life-threatening complications that may result in death or a maternal near-miss, to mild sequelae—and duration. Meanwhile, the relatively minor physical problems that commonly follow childbirth, such as backache, fatigue, vaginal pain, or hemorrhoids, are less studied, although they affect an estimated two-thirds of postpartum women and can incur severe functional limitations [10].

Maternal morbidities affect physical, emotional, economic, and social aspects of women’s lives and influence fetal and infant health and survival. For example, certain complications in pregnancy increase the risk of stillbirth or preterm deliveries, and postpartum complications limit women’s ability to breastfeed, care for, or interact with their infants [11, 12]. Understanding the effects of maternal morbidities on women’s quality of life (QoL) is therefore essential for improving health, well-being, and productivity among childbearing women. The objective of this analysis was to investigate women’s lived experiences with maternal morbidities during pregnancy and postpartum across five LMICs and understand the challenges they present to women’s QoL. Findings will inform development of tailored interventions to improve the QoL of childbearing women in LMICs.

## MATERIALS AND METHODS

### Study design and participants

This qualitative study was conducted at six research sites across five LMICs. Sites were located in Kintampo, Ghana; Kisumu, Kenya; Lusaka, Zambia; Karachi, Pakistan; Vellore, India; and Hodal, India. Data were collected between November 2023 and June 2024, during focus group discussions (FGDs) with pregnant and late postpartum women (6 months to 1 year postpartum) and in-depth interviews (IDIs) with early postpartum women (≤6 weeks postpartum). Eligible participants were women who were either currently pregnant or had delivered in the past 12 months, lived in the catchment area, met the minimum required age (Ghana, Zambia, and Pakistan: 15 years; Kenya and India: 18 years) and provided written informed consent.

Purposive sampling was used to identify potential participants. Recruitment leveraged existing household demographic surveillance systems, antenatal and postnatal care clinics, and the ongoing Pregnancy Risk, Infant Surveillance, and Measurement Alliance (PRISMA) Maternal and Newborn Health cohort study [13]. Efforts were made to recruit evenly across age groups, trimesters, local languages (if multiple), and time since childbirth (“early postpartum” ≤6 weeks and “late postpartum” 6 months to 1 year). Pregnant and late postpartum women participated in FGDs at a healthcare facility or community location; early postpartum women completed an IDI at their home. It was not possible to conduct FGDs with early postpartum women because of cultural practices that discourage women from leaving the home in the early weeks following childbirth. FGDs were stratified by perinatal status (i.e. pregnant or late postpartum) and age group (conducted separately with women <25 years and ≥25 years of age); each contained 8 to 10 women across all three trimesters (if pregnant). Recruitment continued until the target sample size was reached (determined a priori based on budgetary constraints).

Three or four FGDs and eight IDIs were conducted in the local language at each of the six sites. FGDs and IDIs were led by a female trained moderator/interviewer with postgraduate education and experience in qualitative research. The moderator/interviewer was assisted by a research assistant, also female, from the site country, and fluent in the local language and English. In most cases, either the moderator/interviewer and/or the assistant were involved with the PRISMA study and therefore may have been familiar to some participants. During the consent process, the following was stated: “The purpose of this study is to understand mothers’ experience during pregnancy and after childbirth”. Before starting the FGD or IDI, participants were asked to complete a brief sociodemographic and obstetric history questionnaire, which was administered verbally by a member of the research team.

Semi-structured guides for the FGDs and IDIs were developed collaboratively by the research team, with input from investigators at all six study sites and the coordinating site at George Washington University (GWU) (**Supplement 1**). The development of the guides was informed by existing conceptual frameworks of maternal morbidities [14, 15]. The guides included questions about how women in the community care for themselves during pregnancy or postpartum, the main challenges that affect pregnant and/or postpartum women, how these challenges are viewed by family and community members, and how women could be better supported during pregnancy or postpartum. Postpartum women were also asked about their delivery and any challenges or complications experienced during the birth. If a participant indicated experiencing a complication, the interviewer asked additional questions to probe for further details. No repeat interviews were carried out.

The FGD and IDI guides were forward translated (i.e. English to local language) and back translated (i.e. local language to English) per a standardized translation protocol. Minor revisions to some questions were made and additional probes added after beginning data collection to enhance clarity and elicit meaningful responses. Specifically, the FGD guide was updated to include probes based on feedback from the sites about how family and community view physical, emotional, social, and economic challenges during pregnancy and how women seek healthcare while pregnant. The IDI guide was updated to include probes about women’s expectations regarding the birth of their child and how women care for their health postpartum.

FGDs were 60-90 minutes in duration and IDIs were 30-45 minutes. All FGDs and IDIs were recorded and the recordings were transcribed verbatim in the local language. Transcripts were subsequently translated into English, ensuring that the essence was intact. All transcripts were reviewed for completeness, anonymity, and clarity prior to coding and analysis.

### Data Analysis

A collaborative, multi-step approach was used for data analysis. After reviewing the transcripts for familiarization with the data and confirming that saturation had been reached, a subset (2 to 3 FGDs and 1 to 2 IDIs) of transcripts from each site was coded independently by two coders (one from the local site and one from the coordinating site) using an inductive approach, to develop a site-specific codebook. The site-specific codebooks were then merged into a shared cross-site codebook and discussed with all site investigators. All transcripts were then coded independently by two coders in Dedoose™ using the shared cross-site codebook, which was iteratively refined by adding, merging, and reorganizing codes throughout the coding process. After finalizing the shared cross-site codebook, all transcripts were independently reviewed and re-coded by the two coders to ensure alignment with a final shared cross-site codebook. Initial themes and subthemes were identified and subsequently discussed with investigators at all local sites. Participant input into the themes and subthemes identified was not solicited. The themes and subthemes were refined iteratively and collaboratively through discussion with the research sites, after which, representative quotations were selected.

### Ethical considerations

All participants provided written informed consent or assent prior to participating. Study procedures were reviewed and approved or exempted by the Institutional Review Board (IRB) and Ethics Review Committee (ERC) at each participating institution, as follows: The George Washington University, United States (NCR235136); Kintampo Health Research Centre, Ghana (KHRCIEC/2023-32); Society for Applied Studies, India (SAS/ERC/MMS Study/2023); Kenya Medical Research Institute, Kenya (SERU 4882); Aga Khan University, Pakistan (2023-9286-26951); Christian Medical College Vellore, India (IRB Min. No. 15913); University of Zambia, Zambia (Ref No. 4442-2023); and the University of North Carolina at Chapel Hill, United States (Z32301).

## RESULTS

A total of 23 FGDs (4 FGDs per site except Kenya who conducted 3 FGDs) and 48 IDIs (8 IDIs per site) were conducted. Across all sites, 62% of women invited to participate presented for their scheduled session and provided informed consent (overall 253/411; by site: Hodal, India 39/82; Vellore, India 38/77; Ghana 43/58; Pakistan 47/56; Zambia 48/91; Kenya 38/47). Data from 1 postpartum IDI participant (Vellore, India) were excluded during the analysis because the participant had already participated in a FGD during pregnancy.

Participants’ characteristics are summarized in **Table 1**. Most participants were multipara (75%) and married or cohabitating (93%). The mean age of study participants was 26 years (range: 16 to 46 years), with 46% under 25 years. Among pregnant participants, gestational age distribution was skewed toward later pregnancy, with 10% in their first trimester, 32% in their second, and 58% in their third trimester. Occupational trends differed across sites, with >85% of women reporting being housewives or not employed in Pakistan and India, compared to 23% in Ghana, 45% in Kenya, and 71% in Zambia. Overall, the most common occupations were working for a small business or as a business owner (15%) or as a skilled laborer (6%). The reported heads of household also differed regionally, with most reporting husband/partner at the African sites, versus parent/parent-in-law among participants at the Asian sites.

Four overarching themes were identified using thematic analysis: (1) physical and emotional challenges pose a barrier to daily activities; (2) lack of social support detracts from women’s QoL; (3) receiving social support mitigates adverse impacts of pregnancy and postpartum challenges on QoL; and (4) economic challenges exacerbate declines in women’s QoL during pregnancy and postpartum. The overarching themes intersected pregnancy and postpartum and were cross-cutting, irrespective of continent or country. Some differences in minor themes and subthemes were observed across pregnancy and postpartum women and/or across regions or countries.

### Theme 1. Pregnancy and postpartum challenges pose a barrier to daily activities

Physical morbidities, such as pain and fatigue, were a near-universal experience during pregnancy and postpartum and hindered women’s ability to carry out daily activities (**Table 2**). Pain associated with bending and lifting made it difficult for women to complete household work, particularly chores such as washing, sweeping, and fetching water. As one postpartum woman in Vellore, India said, *“Because of my back pain, I can’t bend and do any work.”* Feelings of weakness and fatigue were also commonly reported. For some, this made it impossible to complete tasks that involved heavy lifting, or any tasks at all without taking breaks. A pregnant woman in Lusaka, Zambia said, *“I feel body weakness like I have just woken up. Sometimes I am very weak that I only manage to do laundry once in a week and just some light work. Most of the time I just want to sit and sleep.”* Many women also indicated that they would have liked more help from family members to reduce their workload.

Emotional changes during pregnancy and postpartum also posed a challenge to daily functioning (**Table 2**). Women described a variety of emotional challenges, including mood swings, depression, anger, anxiety, brain fog, and feelings of helplessness. In some cases, this also manifested as physical morbidities. As one pregnant participant in Lusaka, Zambia said, *“I have become very moody and high-tempered, sometimes I feel pain in my heart. I think this is not good for a pregnant woman.”* Some women found it difficult to control their emotions, which made it hard for them to be productive and complete the work expected of them. For example, a postpartum woman in Karachi, Pakistan said, *“When I get angry, then I just go outside. I start to lose consciousness from the anger.”*

### Theme 2. Lack of social support detracts from well-being during pregnancy and postpartum

Women described receiving inadequate support from the baby’s father, as well as from their families and communities (**Table 3**). With regard to the baby’s father, participants explained that they received a lack of support in several areas, which included practical support (e.g. household chores or childcare), emotional support, and financial support. For example, a postpartum woman in Lusaka, Zambia said, *“After delivery, every woman expects their husband to be happy and support them, but this doesn’t happen to everyone, so many women go into postnatal depression because of lack of care from their spouses.”* Participants also expressed that the baby’s father did not recognize or acknowledge their feelings or need for rest when they were physically and or mentally exhausted, which contributed to feeling isolated. As explained by a pregnant woman in Kintampo, Ghana, *“If I say I am tired and cannot anymore, he will not understand and will not give me the needed attention.”*

Insufficient financial support was also widely identified as a key challenge, with the baby’s father sometimes unable or unwilling to contribute to expenses for food, medical care, or basic supplies. As a participant in Kisumu, Kenya said, *“Sometimes when you get pregnant and realize, as a woman, you will go to your husband and tell him that you should start saving and buy the baby’s necessities; but he won’t be even interested in even giving out the support that you would need.”* Women explained that the absence of financial assistance forced women to return to work earlier than expected after childbirth, which exacerbated physical challenges such as back pain and delayed or precluded their recovery.

Some participants described mistreatment from the baby’s father, including being subjected to physical violence, psychological abuse, and withholding of basic necessities, such as food. As a postpartum woman in Lusaka, Zambia said, “I was being beaten by my husband, claiming that I didn’t have respect for him. He would torture me by not providing me with food, and I would just drink water.” Participants’ also described experiences of infidelity and rejection from their baby’s father, which was particularly salient at the Zambia site.

Outside of the home, some women described feeling judged by their community as being lazy when they were weak or unwell, as well as being compared to other pregnant women who did not suffer such symptoms. One pregnant woman in Kintampo, Ghana said, *“I am currently not feeling well but another pregnant woman might be okay so if I show any signs of weakness, people might say I am being lazy.”* Participants explained that community members pass judgment on the way women raise their children and expressed a desire for more empathy and/or assistance from their community. As a pregnant woman in Vellore, India explained *"There’s no one to offer help or even ask if [pregnant women] are tired. No one is there to inquire if they need assistance. Even if there are many people around them, no one cares enough to ask how they feel personally."* A subtheme specific to India and Pakistan, was that participants perceived the medical care they received as inadequate. Women reported that hospital staff were not attentive to their physical and emotional needs during pregnancy or during labor and delivery. In some cases, participants spoke of broader systemic challenges, including lack of nearby clinics and insufficient health care professionals.

A lack of support from family members further detracted from participants’ well-being. For example, participants explained that their family members did not offer to help with household chores. One pregnant woman in Hodal, India said *“We have no choice but to do all the work. Like living alone with my husband, father-in-law, and one and a half-year-old daughter, I have to take care of all of them, clean my house, and make food and I have no one for help. Even if I am on a ventilator, I will be the only one who will do all the work [joked and smiled].”* Participants shared that their family members had unreasonable expectations and pressured them to complete tasks they were unable to do. A minor theme, specific to India and Pakistan, was this criticism and pressure was commonly inflicted by the mother-in-law. As a postpartum woman in Karachi, Pakistan said *“I have to complete my tasks before resting otherwise the family…I can’t just tell them I’m resting now and will serve food later, can I? If I say that, my mother-in-law would take it badly, thinking I’m being lazy or neglectful.”*

One specific source of stress stemmed from societal pressure to have a male child (**Table 3**). Participants described being looked down upon and resented for having a girl and feeling upset as a result. This was especially present in India, where it is illegal to determine the sex of the fetus and female feticide remains a major concern. As one woman in Hodal, India said, “Her husband drinks and beats her up because she gave birth to 5 girls.”

### Theme 3. Receipt of social support mitigates pregnancy and postpartum challenges

Participants shared that receiving support from their families and communities made managing pregnancy and postpartum challenges easier (**Table 4**). For example, women were able to rest and recover when family members helped with household chores and were appreciative of emotional support such as extra care, attention, and pampering. One postpartum woman in Kintampo, Ghana said, *“I live peacefully with the people in my house so I get people to bathe the baby and even if I want, they will want to bathe and massage me. For the baby, they will take very good care of him. They will cook and bathe the baby until you ask them to stop.”* Some family members also provided financial assistance for expenses such as medical costs during pregnancy and buying food and supplies for her recovery after childbirth, which alleviated economic pressures.

Community support further mitigated challenges during pregnancy and postpartum and elicited feelings of belonging. For example, participants explained that their neighbors helped with physical tasks during pregnancy, such as fetching water, and assisted with transportation to the hospital at the time of delivery. As a postpartum woman in Karachi, Pakistan said, *“In my case, my neighbor, although she is not closely related, took care of me just as my sisters would have. She didn’t leave anything lacking.”* Friends also provided support by preparing meals and washing clothes, especially when women did not have that support from their families. One woman in Kintampo, Ghana said, *“When you give birth in the community your loved ones and friends can come and help with cooking, washing clothing, and fetching water for you.”* Women explained that they felt valued by their community when their needs were recognized and prioritized. For example, receiving priority seating on public transportation or at community gatherings provided participants with a sense of comfort and belonging within the community.

### Theme 4. Economic challenges detract from health/well-being in pregnancy and postpartum

Economic hardship was described by participants across all study sites (**Table 5**). Participants explained that they struggled to afford their prenatal and postnatal care, including the transportation costs to travel to and from the healthcare facility. A woman in Lusaka, Zambia said, *“…a pregnant woman is required to carry out some tests, but they don’t have money.”* In some cases, participants explained that the baby’s father or their family had to spend beyond what they could afford to cover the prenatal and delivery expenses, which compromised their financial situation. Food insecurity was also a widespread concern. Participants spoke about not having food at home and relying on the baby’s father to bring them something to eat, with some women forced to resort to begging. As a participant in Lusaka, Zambia described, *“I have seen some women who have not fully recovered from childbirth, going round in the community begging for food with a very small baby, because they had no food at home.”*

While financially necessary, returning to work or seeking employment after childbirth was complicated by physical challenges and lack of childcare. Many women described being physically exhausted and without the strength to work. One woman in Vellore, India said, *“I wanted to return to the office and thought I could work, but since I can’t sit for more than a few hours, I don’t know how I would manage in the office.”* Even among mothers who were physically capable, a lack of caretakers or financial ability to pay for childcare precluded them from seeking employment. Those who did return to work often did so prematurely, compromising their health to support themselves and their baby, and often against the wishes of their families. As described by a woman in Hodal, India: *“After having a baby, women often leave their job. Their family tells them that they are incapable of taking care of the baby. Therefore, they tell women to choose one between their home and career. I have seen many cases in our village like this.”*

## DISCUSSION

This study aimed to understand women’s lived experiences of maternal morbidities during pregnancy and postpartum and their impacts on QoL, across five countries in sub-Saharan Africa and South Asia. Participants described experiencing a range of physical and emotional challenges, many of which prevented them from carrying out daily activities such as completing household chores, obtaining food and necessities, and/or maintaining self-care. Consistent with existing literature, participants reported severe backaches and musculoskeletal strain, often prompting them to avoid or modify strenuous tasks like fetching water g [16]. Fatigue, driven by hormonal changes and sleep disruptions, emerged as a major barrier, aligning with previous studies on postpartum exhaustion [17]. This reflects how sociocultural expectations intensify the burden on women, despite their physical limitations. Financial barriers, limited social support (particularly from partners), and sociocultural expectations further exacerbated these difficulties.

Nevertheless, family and community support emerged as critical mitigating factors, underscoring the importance of informal networks in improving maternal well-being. The importance of receiving social support during the perinatal period has been described previously [18, 19]. Poor social support during pregnancy and postpartum is associated with perinatal depression, post-traumatic stress, and other mental health concerns [20]. A systematic review on women’s experiences of social support during pregnancy found that pregnant women lacking emotional connection and reassurance from their partners were more likely to experience anxiety and depression [18]. Previous research in India and Pakistan showed that women with perinatal depression had lower scores on QoL domains [21]. Postpartum depression has also been linked to poor physical health and insomnia, further exacerbating the functional impacts of maternal morbidities [10, 22, 23]. Conversely, several prior studies demonstrate that social support is positively associated with QoL [24, 25] and receiving emotional support from family improves maternal mental health outcomes [18, 26, 27].

The extent to which women negatively described interactions with the baby’s father and/or their mother-in-law was notable, though not altogether surprising. In sub-Saharan Africa, cultural norms discourage male involvement in caregiving roles [26, 28–30] and similar trends have been observed in parts of Asia, where patriarchal norms restrict male engagement in maternal care [31, 32]. The cultural acceptance of infidelity, especially in sub-Saharan Africa, further fosters feelings of abandonment and neglect, which heightens the risk of postpartum depression [32].

Mistreatment from partners—including intimate-partner violence, rejection, and infidelity—has been previously shown to worsen maternal physical and mental health outcomes [33–35]. Intimate-partner violence has been associated with adverse birth outcomes including preterm birth and low birth weight [36]. Community and family pressures, such as the stigmatization of single mothers and societal expectations regarding the sex of the baby, further exacerbate feelings of social isolation and emotional challenges, which are disproportionately prevalent in patriarchal societies [35, 37, 38]. Importantly, as research by Cumber and colleagues (2024) reveals, fathers’ motivations to be more supportive may also be hindered by cultural stigma, financial barriers, or exclusion from maternity services [39]. This calls for intentional involvement of expectant fathers in prenatal care to foster their participation.

Women in the present study cited economic challenges as a barrier to accessing care or purchasing necessities, such as transportation, food, and clothing for themselves and/or their baby. Existing literature emphasizes how financial insecurity—particularly a lack of financial contributions from the baby’s father—can severely restrict access to prenatal care and nutritious food [18, 40, 41]. A lack of prenatal care worsens maternal morbidities [41, 42], whereas timely prenatal care reduces pregnancy-related complications and adverse fetal outcomes [43–46]. Maternal malnutrition during pregnancy is further tied to maternal morbidities [47–49] and nutritional deficiencies postpartum increase women’s vulnerability to illness, reducing their capacity to care for themselves and their newborns [50]. Economic hardships also presented women with stressful decisions; for example, postpartum participants described needing to choose between earning money and caring for their child, which further restricted their financial independence [51–54]. This is particularly relevant in patriarchal societies where traditional gender roles place the burden of childcare on women.

Key strengths of the study include data collection across five LMICs on two continents, which allowed us to capture and compare women’s experiences during pregnancy and postpartum in different cultural contexts. Another strength of the study was the enrollment of women across trimesters and at varying time points postpartum, which allowed us to more comprehensively assess women’s experiences of maternal morbidities and their impacts on QoL. Several limitations also warrant consideration. Considering the hierarchical nature of study contexts, participants may have responded in a socially desirable manner. The added group dynamic of the FGDs or lack of privacy of IDIs—given they occurred within the participants’ homes—may have further discouraged women from engaging in open dialogue.

## CONCLUSIONS

These results demonstrate that maternal morbidities have wide-reaching impacts on women’s quality of life during pregnancy and postpartum, particularly in carrying out household chores, obtaining food and other necessities, and maintaining self-care. Consistent with previous research, social support was a vital factor in mitigating declines in health and well-being associated with childbearing. Our findings suggest that interventions involving fathers and family members in maternity care and educating them about the challenges and limitations women face during pregnancy and postpartum are needed, as well as integrating QoL assessments in routine medical care. Targeted strategies to increase access to physical and emotional support for pregnant and postpartum women, promote economic empowerment, and provide financial support during the perinatal period are also critical to support women’s health and well-being during pregnancy and postpartum.

## AUTHORS’ CONTRIBUTIONS

Conceptualization: SGB, JK, KM, ERS, ACS, and NJV. Funding acquisition: VA, KPA, AGC, ZH, MPK, SM, WM, IN, ERS, and MBS. Design of methodology: SGB, JK, KM, ERS, ACS, and NJV. Investigation—data collection: MA, PA, ME, EBK, AK, DL, GPM, PO, GO, WP, JOP, NS, and CT. Project administration: SGB and NJV. Software—code development: ACS and NJV. Formal analysis: MA, PA, BMB, SB, EBK, ME, GDG, AK, MM, PO, GO, PP, WP, JOP, and NJV. Supervision: VA, KPA, AGC, ZH, MPK, SM, WKM, IN, NS, ERS, MBS, ACS, and CT. Writing—original draft preparation: MA, SGB, BMB, GDG, MM, KO, PP, JOP, MBS, NJV, and ME. Writing— review and editing: MA, SGB, AGC, ZH, JK, EBK, SM, WKM, PO, MBS, ERS, and ACS.

## Data Availability

Transcript data will not be made publicly available given that it contains private and confidential information. However, all data collection tools and protocols will be made available on Open Science Foundation under the PRISMA Consortium (osf.io/qckyt).

## ACKNOWLEDGEMENTS

This study would not be possible without the support from the Bill & Melinda Gates Foundation, specifically from Drs. Laura Lamberti and Sun-Eun Lee. The authors would also like to acknowledge the community members, especially pregnant and postpartum women, whose generous participation helped to answer our research questions.

